# The Effect of AI on the Radiologist Workforce: A Task-Based Analysis

**DOI:** 10.64898/2025.12.20.25342714

**Authors:** Curtis P. Langlotz

## Abstract

**Background:** The effect of AI algorithms on the radiology workforce has been a subject of commentary and controversy. There is now sufficient published evidence to support a quantitative task-based analysis to predict these effects.

**Purpose:** To construct a quantitative, task-based model to predict the effect of AI on the radiology workforce using the best available evidence.

**Materials and Methods:** We reviewed the literature to establish the tasks on which radiologists spend their time. We then developed categories of AI applications that could affect these tasks. We used published evidence to estimate the effect of each AI application on each radiology task using a 5-year time horizon. When published evidence was unavailable, we used our own judgment.

**Results:** The model projects a 33% reduction in hours worked by radiologists in 5 years, with a range of 14% to 49%. The main effects are due to radiology report drafting for all modalities and study delegation for radiography and mammography.

**Conclusion:** AI applications likely will cause a significant decrease in radiologist hours worked.. Given the relatively static radiology workforce and the continued growth in imaging volumes, radiologist job loss is unlikely for the foreseeable future.

## Introduction

Ten years ago, many experts predicted machine learning would displace much of the work of radiologists [1–5]. Since then, a substantial radiology AI industry has arisen, with more than 100 AI companies exhibiting at the most recent radiology professional meeting [6] and more than three-quarters of FDA cleared AI/ML software devices targeting radiology [7].

As radiologists gain experience with the actual use of AI systems, extensive commentary has questioned how AI will affect the radiology workforce [8–13]. The answer is of clear interest not only to radiologists, but also to many other stakeholders, including medical students choosing a medical specialty, radiologists-in-training who will be affected by these developments throughout their careers, other medical professionals who may see AI’s effects in radiology as a leading indicator of how AI may affect their workforce, and even to professionals in other industries, who see radiologists as an example of a highly-skilled worker whose livelihood may be affected by AI.

The growing scientific literature regarding the effect of AI algorithms on radiology tasks enables for the first time specific quantitative predictions about the effect of AI on the radiology work force. This motivated us to conduct a task-based analysis to project the effects of AI on the radiology work force using the best available evidence.

## Materials and Methods

We reviewed the literature to identify the tasks on which radiologists spend their time. We then reviewed the literature and identified 15 AI applications that may affect the time radiologists spend with each task. Next, we collated the best published evidence for how each AI application will affect each task. We calculated the total effect of AI or radiologist hours worked in the base case and using a lower and upper bound.

### Tasks on Which Radiologists Spend Their Time

Labor economists argue that task-level analysis is the best method to quantify the impact of AI on any profession [14]. In labor economics, a task is a unit of work activity that produces output. Two observational studies have identified and measured the tasks on which radiologists spend their time. Dhanoa et al [15] observed 14 radiologists across 3 Canadian hospitals, one academic tertiary care and two community-based practices, quantifying time spent performing the task categories they defined. Schemmel et al [16] conducted a similar focusing on a neuroradiology fellow at an academic hospital.

We analyzed and categorized the tasks measured, then averaged the percentage of time spent in each category between the two studies. (See Supplementary Materials). These calculations defined the time spent on specific radiologist tasks: Perform and interpret the study, 66.7%, protocoling, 5.5%; communicate with technologist or nurse, 3.9%; communicate with radiologist, 3.7%; communicate with provider, 3.3%; Answer order question, 0.7%; communicate with patient 0.2%; other, 6.8%; personal, 9.1%.

Because the effect of AI tools varies across imaging modalities, we estimated the fraction of time radiologists spend on interpreting studies from each modality based on the number of studies performed each year in the US [17–22], the percentage of those studies interpreted by radiologists, and a work RVU adjustment commonly used by radiology practice to measure productivity [23]. (See Supplementary Materials). Since these data were weighted toward academic practice, we also examined how our results would change if image interpretation consumed 80 percent of radiologists’ time, with a compensatory reduction in time consumed by non-interpretative tasks. Figure 1 shows the percent time radiologists spend on job tasks.

**Figure 1.**
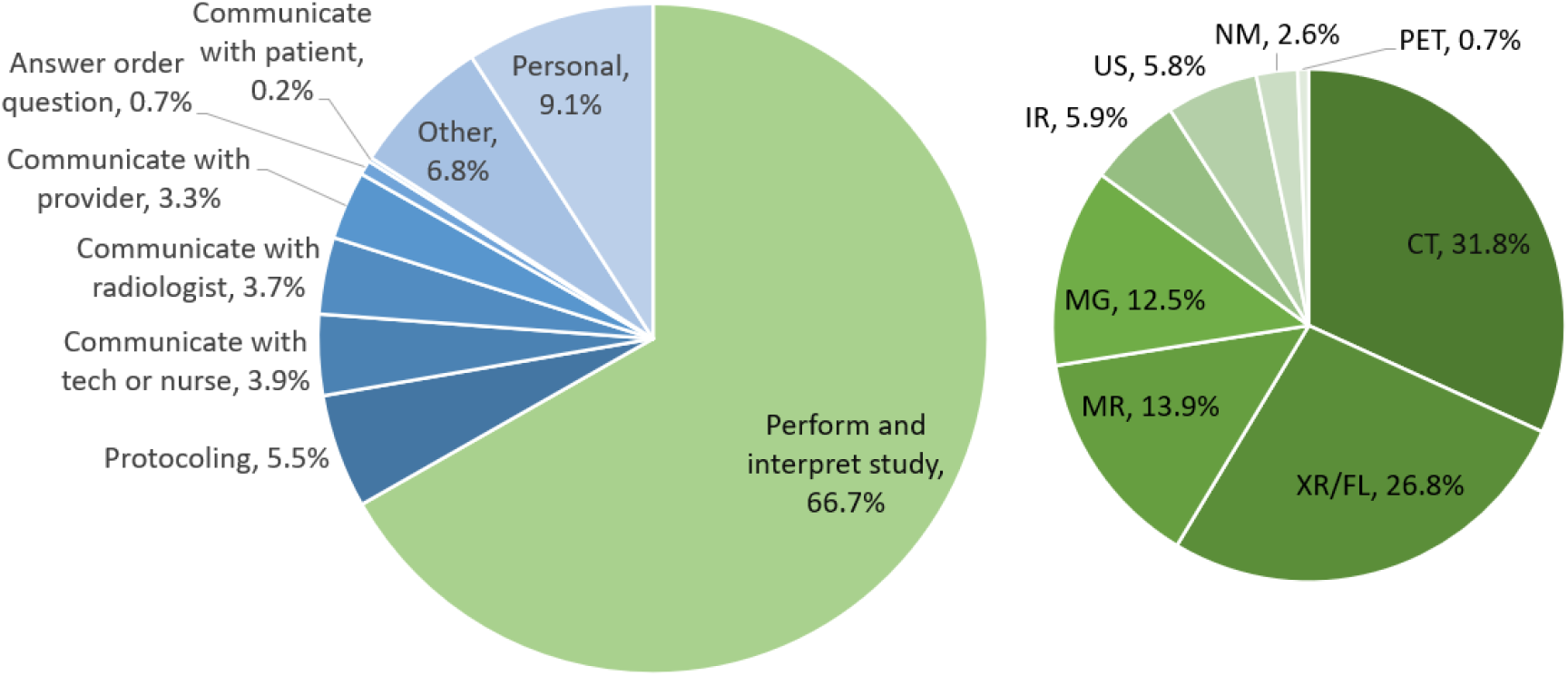
A breakdown of how radiologists spend their time on various tasks, as well as adjusted interpretation times from U.S. federal data.

### Task-Based Analysis of AI Effects

To estimate how AI would affect various radiologist tasks, we reviewed three main sources of information: the CMS Medicare NCCI Coding Policy Manual [24], the Department of Labor O*NET Online database [25], and the published biomedical literature [26].

We adapted and unified these procedure-related work processes (See Supplementary Materials) to assemble categories of AI applications that may affect these processes. The AI applications and their definitions are shown below, grouped by pre-procedure, intra-procedure, and post-procedure applications:

### Pre-procedure applications of AI to radiology

- **Order entry decision support for radiology test selection**. A decision is made whether a test is appropriate based on the patient’s clinical history and current symptoms.
- **Review of patient history**. The patient’s medical history and the reason for the study are reviewed to guide the performance and interpretation of the study.
- **Select and tailor the imaging protocol**. An appropriate imaging protocol is selected based on the information in the patient’s medical record and the reason for the study.

### Intra-procedure applications of AI to radiology

- **Image reconstruction**. The raw data is converted from detected signals into a visible image. This conversion typically occurs within the imaging device itself or using specialized software.

### Post-procedure applications of AI to radiology

- **Imaging study delegation**. Imaging studies are analyzed to identify those that do not require human interpretation.
- **Imaging study triage**. Imaging studies likely to have significant positive findings are identified. Flagged studies are prioritized on the radiologist’s work list so they can be interpreted first.
- **Abnormality detection**. Areas on the imaging study that are likely abnormal are identified, typically by using a heat map, a marker, or other identifier, to call attention to these areas.
- **Disease classification**. Based on the imaging findings, a ranked list of diagnostic possibilities is assembled.
- **Information extraction**. Quantitative information is extracted from the images, such as the diameter of the aorta or the amount of coronary calcium present on a chest CT.
- **Radiology report drafting**. Language models or other documentation tools are used to produce a draft report that describes the findings on the imaging study.
- **Non-routine communication**. Critical or unexpected findings are urgently communicated with the care team so action can be taken.
- **Explain radiology reports**. Language models explain complex terminology found in radiology reports to improve the understanding of clinicians and patients.

### Effect of AI on Radiology Processes

We then considered the magnitude and direction of the effect of each AI application (e.g., radiology report drafting) on each task (e.g., interpretation and reporting). For each interaction of application and task we estimated a base case effect, as well as a lower and upper bound. We assumed that the effects of each AI application on each task were independent. For example, a 3% reduction in imaging volume due to order entry decision support is independent of the effect of report drafting, which would improve efficiency on the remaining 97% of orders. Therefore, we multiplied the percentage effects of each AI application on each radiology task to arrive at the overall reduction in hours needed to accomplish that task.

We considered the implications of adopting the innovations that are available today (e.g., detection and triage), as well as those applications that are foreseeable within a 5 -year time period based on developments in the laboratory and the published literature (e.g., automated report drafting). These modeling assumptions are described below and summarized in Table 1.

**Table 1.** A summary of model assumptions. Numbers in red indicated situations in which AI may increase the work of radiologists.

| AI Product | ↑↓ | Task Affected | Base Case | Range |
| --- | --- | --- | --- | --- |
| AI for order-entry decision support | ↓ | volume (CT/MR/NM/US/PET) | 3% | 0% - 6% |
| AI to summarize the medical record | ↓ | interpretation time | 2% | 0% - 4% |
| AI to summarize the medical record | ↓ | communicate with tech, provider | 30% | 10% - 50% |
| Automated protocoling systems | ↓ | protocoling time | 60% | 30% - 70% |
| Image reconstruction | ↓↑ | Interpretation time (CT, MR, PET) | 0% | 1% - +5% |
| Imaging study delegation | ↓ | need for a human in the loop (MG) | 50% | 30% - 60% |
| Imaging study delegation | ↓ | need for a human in the loop (XR) | 40% | 30% - 50% |
| Imaging study delegation | ↓ | need for a human in the loop (other) | 3% | 1% - 5% |
| Detection algorithms | ↓ | interpretation time (XR) | 5% | 0% - 15% |
| Detection algorithms | ↓↑ | interpretation time (other) | 0% | 5% - +5% |
| Imaging study triage | ↓ | interpretation time | 1% | 0% - 5% |
| Classification algorithms | ↓ | interpretation time | 1% | 0% - 5% |
| Opportunistic screening | ↑ | interpretation time (CT, XR) | +2% | 0% - +5% |
| Radiology report drafting | ↓ | interpretation time | 20% | 10% - 30% |
| Nonroutine communication | ↓ | communicate with provider | 30% | 20% - 50% |
| Radiology report explanation | ↓ | communicate with patients | 30% | 10% - 50% |

### Effect of AI on Pre-procedure Applications in Radiology

- **Order-entry decision support for radiology test selection**. Evidence is mixed regarding whether order-entry decision support reduces diagnostic imaging volume or improves the appropriateness of test ordering [27–33]. Despite past federally mandated implementation of these tools, imaging volume continued to increase. Decision support using machine learning could be more accurate and therefore have stronger effects [34], but there has not yet been an assessment of how these newer systems will affect order volume. We assumed a base case 3% reduction in advanced imaging orders (CT/MR/NM/US/PET) with a range of 0-6%.
- **Review of patient history**. Several laboratories and commercial entities are building AI tools to summarize a patient’s history from the electronic medical record [35,36]. An AI summarization system could reduce the time radiologists spend searching for relevant clinical history in the medical record, particularly for complex patients. There is no published data on how these systems affect radiologist productivity. We assumed a base case 2% reduction in interpretation time for all studies with a range of 0-4%. We also assumed that these systems would reduce the need for communication between radiologists, technologists, and referrers by 30%, with a range of 10-50%.
- **Select and tailor the imaging protocol**. Recent work shows that 69% of studies could be protocoled automatically with 95% accuracy [37]. These new systems could substantially reduce the time radiologists spend selecting imaging protocols for CT and MR. We assumed these systems would reduce protocoling time by 60%, with a range of 30-70%.

### Effect of AI on Intra-procedure Applications in Radiology

- **Image study reconstruction**. Physics-based AI reconstruction methods can acquire images more efficiently than conventional methods and can create images with comparable or slightly lower quality [38–42]. Lower image quality may decrease productivity. In the base case, we assumed that these tools would have no appreciable effect on radiologist productivity, with a range of 1% reduction in interpretation time to a 5% increase in interpretation time.

### Effect of AI on Post-procedure Applications in Radiology

- **Imaging study delegation**. Several studies have shown that a subset of radiology studies can be identified by AI that are almost certainly normal and require no further human review, including up to 50% of screening mammograms [43,44] and up to 40% of outpatient radiographs [45,46]. When combined with human review of the remaining studies, these systems can provide equivalent or improved accuracy compared to a human-only system. More evidence is needed, but these systems could have a large impact on radiologist workload in selected subspecialty areas. We assumed that image delegation would affect time spent interpreting mammograms by 50%, with a range of 30-60%. We assumed the effect on radiography would be 40%, with a range of 30-50%. Although speculative, we assumed a 3% delegation of all other modalities, with a range of 1-5%.
- **Imaging study triage**. Sorting cases by complexity may have a small beneficial effect on radiologist productivity [47]. Otherwise, these tools have no appreciable effect on radiologist productivity because the same number of studies must be interpreted in a different order. We assumed a 1% improvement in interpretation time, with a range of 0-5%.
- **Abnormality detection**. Detection systems may increase efficiency of “needle-in-a-haystack” tasks, such as detecting lung nodules, but may simultaneously decrease radiologists’ productivity by adding the need to adjudicate false positive detections. There is weak evidence for improved productivity when using AI on chest radiographs [48–50], with no evidence regarding other study types. For radiography we assumed a base case of 5% increased productivity, with a range of 0-15%. For other study types, we assumed a base case of no time savings, with a range of 5% savings to 5% increase in time spent.
- **Disease classification**. Differential diagnosis tools may improve efficiency by helping radiologists quickly resolve diagnostic dilemmas, especially for abnormalities outside a radiologist’s subspecialty, such as a lesion seen in the humerus on a chest radiograph. There is no published evidence on the effect of diagnostic decision support on radiologist efficiency. We assumed a base case of a 1% time savings, with a range of 0-5%.
- **Opportunistic screening**. There is substantial and growing literature on AI algorithms that detect abnormalities or quantify findings that radiologists generally don’t extract from images, such as coronary calcium on a non-gated chest CT [51], sarcopenia on an abdomen CT [52], and bone density on chest radiographs [53]. These systems may decrease productivity because radiologists must validate and report the additional findings. We assumed a base case increase in work by 2%, with a range of 0-5%.
- **Radiology report drafting**. Large productivity gains could be achieved if reports are drafted automatically for review and signature. These tools can serve a function like a trainee in practices without trainees [54]. Evidence in the literature cites a 15% productivity increase for radiography [55]. The improved productivity likely would be greater for cross-sectional imaging. We assumed a base case of 20% improvement in productivity, with a range of 10-30%.
- **Non-routine communication**. Assuring that urgent or unexpected results reach the care team in a timely manner can improve the quality of care. Some non-AI solutions to this problem already exist [56]. There is little work on AI-based solutions [57] and no published evidence regarding the effect these systems have on radiologist productivity. We assumed these systems could result in a 30% reduction in radiologist time spent communicating with referring clinicians, with a range of 20-50%.
- **Explain radiology reports to patients**. Vendor products [58,59] and research systems [60] have been developed to explain the complex terms in radiology reports to patients. There is no published evidence on the effect of these communication systems on radiologist productivity. Future work may show these systems reduce patient calls or messages. We assumed these systems reduced the need for patient communications by 30%, with a range of 10-50%.

## Results

We have conducted a task-based analysis to quantify the effect of AI on the radiology workforce over the next 5 years using the best available published evidence. The results are shown in Figure 2. The analysis shows that AI likely will cause an up to 33% reduction in the need for radiologists over the next 5 years, with a range of 14-49%. Our analysis shows that AI has widely varying effects across tasks. Two areas have a strongly positive ROI for radiologists: report drafting, due to its beneficial effect on productivity, and study delegation strategies that reduce volume by placing radiologists out of the loop. On the other hand, opportunistic screening may add to the work of radiologists, who must document additional findings.

**Figure 2.**
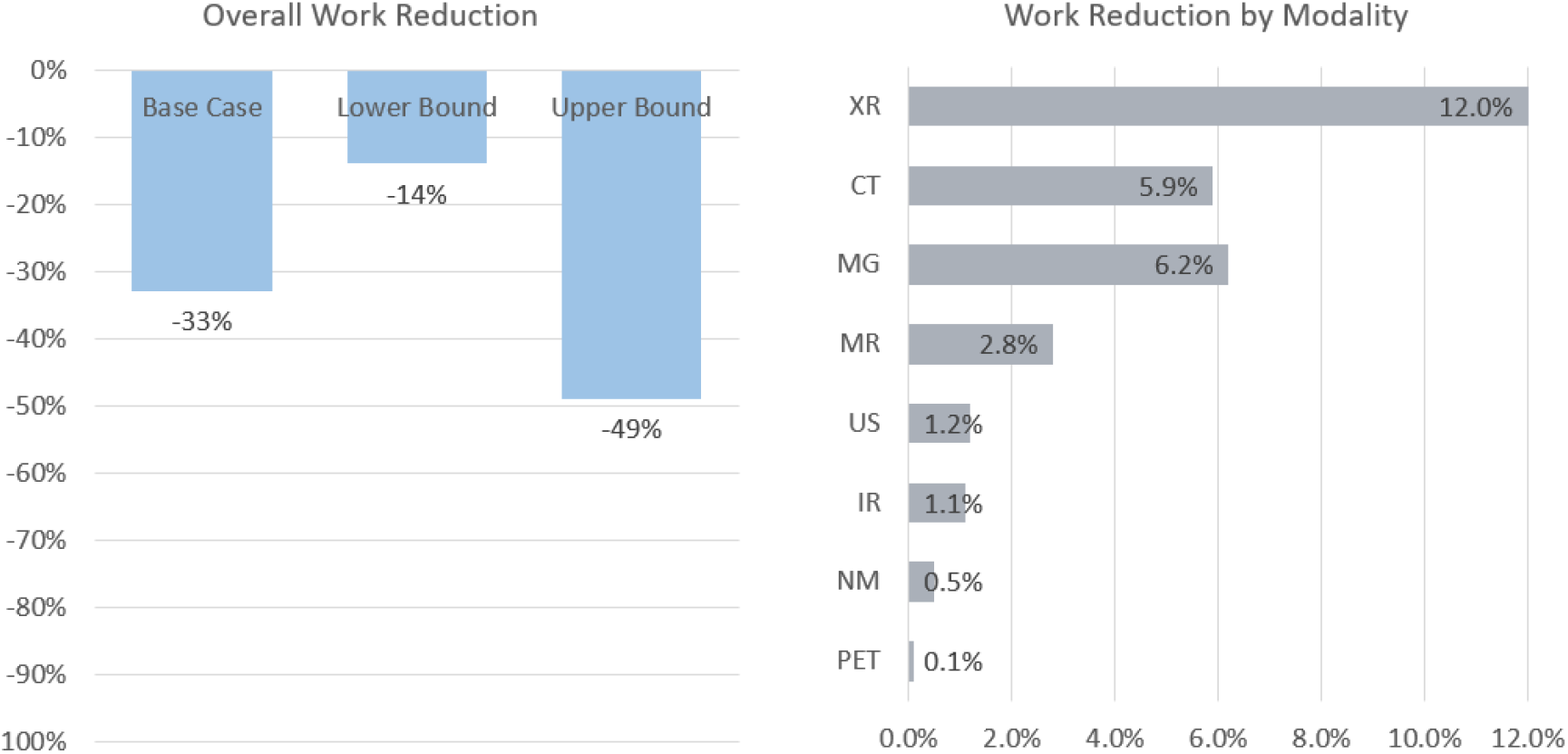
Results of the prediction model. (a) The model predicts a reduction in time worked of 33% for the base case, with an upper bound of 49% and a lower bound of 14%. (b) The main effects are a reduction in time worked to interpret radiography, CT, and mammography, due to the effects of image study delegation and report drafting.

CT, radiography, and mammography experience the greatest effects, primarily because most interpretation and reporting time is spent with CT and radiography, leading to a strong effect on these modalities by radiology report drafting. Mammography and radiography also are strongly affected by imaging study delegation.

Because our estimates of time on task were likely biased toward academic practice, we conducted an analysis that assumed interpretation and reporting tasks consumed 80% of radiologists’ time, with the remaining tasks scaled to consume 20%. The results showed a reduction in radiologist hours worked of 35%, indicating the model is relatively robust to modest changes in task distribution.

## Discussion

These results must be interpreted in the context of the steady growth in imaging volumes, with a near doubling of the volume of cross-sectional studies over the past decade [61]. This trend likely will continue [62]. The projected reduction in hours worked by radiologists is likely to be more than offset by the growth in imaging volumes over the same period, given the relatively static number of radiologists over the same period [63]. This suggests AI may cause shifts among radiologist tasks, rather than a reduction in the need for radiologists.

This study has several limitations. There are no reliable measurements of the effects of AI on many radiology tasks, so many of the model assumptions were estimated from the author’s experience and observations.

We modeled the effects of AI by modality. We did not model the differing effects of AI applications across modality subtypes, such as chest vs. abdomen CT, screening vs. diagnostic mammography, and inpatient vs. outpatient radiography. We considered these factors when we estimated base model assumptions and ranges.

Our analysis lumped procedural components of work with interpretation and reporting. There is limited data on the portion of time procedural specialists spend with interpretation and reporting. We considered this limitation when we developed our initial model assumptions.

The published time and motion studies we used to estimate how radiologists spend their time were short in duration and were weighted towards academic practice. There is likely a high variability in how radiologists spend their time across practices and across subspecialties.

The projected workforce effects of AI likely represent the upper end of a range. Although we have assumed a 5-year timeframe, implementations will occur in phases. Organizational investment in AI will be determined by resources available and the return on investment to each organization. It is unlikely that all organizations will adopt all these algorithms. Some changes, such as the delegation of normal studies to AI-only interpretation, may face significant resistance to adoption.

The analysis is U.S.-centric. Other countries will have different distribution of imaging modalities, and different practices, such as double reading of mammograms, that will lead to different workforce effects.

We did not explicitly consider the productivity effects of new AI governance tasks such as the monitoring and oversight of deployed AI algorithms.

Our conclusions about the overall effect on the radiology workforce depend on the future growth of imaging volumes, which was not explicitly modeled and may not continue.

## Conclusion

We constructed a quantitative, task-based model to predict the effect of AI on the radiology workforce. Our model suggests that radiologist job loss due to AI is unlikely in the next 5 years. The productivity gains from AI are likely to be offset by continued growth in imaging volumes, causing continued growth in the need for radiologists, albeit at a slower rate. Currently approved AI detection and triage algorithms are likely to have only a minimal effect on radiology workload, with tools for study delegation and report drafting having stronger effects, primarily on CT, radiography, and mammography. Overall, these changes may improve the quality of life of radiologists by flattening the growth in their workload and by performing tasks that radiologists consider less interesting, such as lesion measurement and “needle-in-a-haystack” tasks. Because there is a shortage of radiologists in most areas, some decrease in volume (or decrease in growth rate) might be welcomed by radiologists. These results suggest that the effect of AI will be similar to prior waves of innovation in radiology, such as picture archiving and communication systems, which substantially improved productivity but did not displace radiologists.

## Data Availability

All data produced in the present work are contained in the manuscript.

## Acknowledgments

This work was supported in part by the National Institute of Biomedical Imaging and Bioengineering (NIBIB) of the National Institutes of Health under contract 75N92020C00021 and through The Advanced Research Projects Agency for Health (ARPA-H).

