## Supplemental Materials for "The Effect of AI on the Radiologist Workforce: A Task-Based Analysis"

**Supplementary Materials**

### How Radiologists Spend Their Time

We used the results of [15] and [16] to estimate how radiologists spend their time. We analyzed and categorized the tasks measured in each study, then averaged the percent time spent in each category. (See Supplementary Table 1.)

**Supplementary Table 1:** Calculation of the average time spent in each task category, using data from [15] and [16]. (Percentage may not sum to 100% due to rounding.)

| Task Category | Schemmel et al Task | Percent of Time | Dhanoa et al Task | Percent of Time | Average |
|---|---|---|---|---|---|
| | Image interpretation | 37.7% | Interpret images | 36.4% | |
| | Staff-out | 16.1% | Teaching | 14.6% | |
| | Teaching | 1.0% | | | |
| Image interpretation | | 54.8% | | 51.0% | 52.9% |
| | Procedures | 16.7% | Image-guided procedures | 10.9% | |
| Procedures | | 16.7% | | 10.9% | 13.8% |
| | Technologist protocol request | 1.4% | Protocol studies | 3.5% | |
| | Protocoling studies | 6.2% | | | |
| Protocoling | | 7.5% | | 3.5% | 5.5% |
| | Technologist image check | 0.9% | Phone consult, technologist or nurse | 1.1% | |
| | Technologist procedure question | 0.6% | Office consult, technologist or nurse | 4.9% | |
| | Technologist procedure question | 0.2% | | | |
| | Technologist other | 0.1% | | | |
| Communicate with technologist or nurse | | 1.8% | | 6.0% | 3.9% |
| | Interradiologist consult | 0.3% | Office consult, radiologist | 3.1% | |
| | Interradiologist consult | 4.0% | | | |
| Communicate with radiologist | | 4.3% | | 3.1% | 3.7% |
| | Provider study review | 1.9% | Phone consult, physician | 1.2% | |
| | Provider other | 0.2% | Office consult, | 2.0% | |

| | | | | | |
|---|---|---|---|---|---|
| | | | physician | | |
| | Provider study review | 1.2% | | | |
| | Provider other | 0.2% | | | |
| Communicate with provider | | 3.4% | | 3.2% | 3.3% |
| | Provider order question | 1.3% | | | |
| Answer order question | | 1.3% | | 0.0% | 0.7% |
| | Procedure/study consent | 0.4% | | | |
| Communicate with patient | | 0.4% | | 0.0% | 0.2% |
| | Other | 0.2% | Other | 4.4% | |
| | Meetings | 3.1% | Meeting, out of office | 5.5% | |
| | | | Technical downtime | 0.3% | |
| Other | | 3.3% | | 10.2% | 6.8% |
| | Personal: out of room | 4.2% | Personal | 7.1% | |
| | Personal: in room | 1.8% | Meals | 5.2% | |
| Personal | | 5.9% | | 12.3% | 9.1% |
| GRAND TOTAL | | 99.6% | | 100.2% | 99.9% |

We estimated the number of hours worked in each radiology specialty using data on the number of studies interpreted per year in the U.S., the percent interpreted by radiologists, and the mean adjusted work RVU per study. Supplementary Table 2 shows the results of that analysis.

Supplementary Table 2: Calculating the number of hours worked by modality.

| **Imaging modality** | **Studies per year (millions)** | **Interpreted by rads (%)** | **Interpreted by rads (millions)** | **Mean wRVU per study** | **Adjusted AAARAD work RVUs** | **Relative interpretation time index** | **% time on each modality** |
|---|---|---|---|---|---|---|---|
| **Computed tomography (CT)** | 93.0 | 97.3% | 90.5 | 1.41 | 0.45 | 57.4 | 31.8% |
| **Radiography and fluoroscopy (XR/FL)** | 275.0 | 76.6% | 210.7 | 0.23 | 1.0 | 48.4 | 26.8% |
| **Magnetic resonance imaging (MRI)** | 34.7 | 91.0% | 31.6 | 1.77 | 0.45 | 25.2 | 13.9% |
| **Mammography (MG)** | 43.0 | 99.0% | 42.6 | 0.53 | 1.0 | 22.6 | 12.5% |
| **Interventional (IR)** | 7.0 | 85.0% | 6.0 | 4.00 | 0.45 | 10.7 | 5.9% |
| **Ultrasonography (US)** | 37.6 | 33.9% | 12.7 | 0.82 | 1.0 | 10.5 | 5.8% |
| **Nuclear and molecular imaging (NM)** | 9.9 | 50.9% | 5.0 | 0.80 | 1.15 | 4.6 | 2.6% |
| **Positron emission tomography (PET)** | 2.8 | 50.9% | 1.4 | 2.10 | 0.45 | 1.3 | 0.7% |
| **Total** | 503.0 | | 400.4 | | 1.0 | 180.7 | 100.0% |

**How AI Affects Radiologist Tasks**

The Center for Medicare and Medicaid Services (CMS) describes what radiologists are paid for. It also provides useful grouping of radiology tasks into pre-procedure, intra-procedure, and post-procedure categories. We will use this framework to organize the potential effects of AI on radiology tasks.

The O*NET program, sponsored by the U.S. Department of Labor, is the primary source of reliable occupation information in the U.S, containing standardized skills, competencies, and occupational requirements for nearly 1,000 occupations. This information is gathered from a variety of sources, including occupational experts, workers, business establishments, job analytic databases, and professional associations. As we collated these task lists, we grouped similar activities and sorted in chronological order. In the biomedical literature, we found one reference [26] that listed radiology tasks to which machine learning applications are likely to apply. We used our own judgment to fill any gaps in how radiology AI applications may affect radiology tasks. The result is shown in Supplementary Table 3.

Supplementary Table 3: Aligning the effects of AI on the radiology workforce from three sources.

| | Chapter IX of the CMS Coding Policy Manual | Radiologists in Department of Labor O*NET Database | Table 2 in Choy et al. |
|---|---|---|---|
| Pre-procedure work | • reviewing | • counsel patients and | • order scheduling |

| | | | |
|---|---|---|---|
| | indications and clinical history<br>• selecting and tailoring the imaging<br>• consulting the referring clinician or patient if necessary | other physicians to explain the processes, risks, benefits, or alternative treatments<br>• Obtain patients' histories from electronic records, patient interviews, dictated reports, or by communicating with referring clinicians<br>• Direct technologists or technicians regarding desired dosages, techniques, positions, and projections<br>• Instruct radiologic staff in desired techniques, positions, or projections | and patient screening<br>• automated clinical decision support<br>• automated exam protocoling |
| Intra-procedure work | • real-time decision-making regarding modification or extension of the imaging acquisition<br>• oversight of contrast administration and management of contrast reactions<br>• radiation or MR safety monitoring if necessary | • Check and approve the quality of diagnostic images before patients are discharged<br>• Review images and information using picture archiving or communications systems<br>• Recognize or treat complications during and after procedures<br>• Develop or monitor procedures to ensure adequate quality control of images | • image acquisition |
| Post-procedure work | • a detailed examination of the images<br>• comparison with prior studies.<br>• post-processing of images<br>• creation and finalization of a complete report<br>• non-routine | • Interpret the outcomes of diagnostic imaging procedures<br>• Prepare comprehensive interpretive reports of findings<br>• Communicate examination results or diagnostic information | • automated detection<br>• automated interpretation<br>• image management and display<br>• image post-processing<br>• quality analytics<br>• automated dose |

| | | | |
|---|---|---|---|
| | communication of any urgent or unexpected findings<br>• attestation of quality measurements<br>• participation in peer review and other quality improvement processes. | to referring physicians, patients, or families<br>• Participate in quality improvement activities including discussions of areas where risk of error is high | estimation<br>• radiology reporting and analytics<br>• correlation and integration of imaging with other data sources. |
| Other (not directly procedure related) | | • Establish or enforce standards for protection of patients or personnel.<br>• Participate in continuing education activities to maintain and develop expertise<br>• Teach at graduate educational level<br>• Establish and enforce radiation protection standards for patients and staff | |
